# Methodological guidelines for circadian modeling of Daylight Saving Time: application to the United States

**DOI:** 10.64898/2026.06.17.26355889

**Authors:** José María Martín-Olalla, Jorge Mira

## Abstract

Modeling the circadian impact of seasonal clock changing requires precise synchronization between solar and social time. This report critiques a recent study that associated disease prevalence in the United States with seasonal clock exposure. We identify a fundamental computational error in which a sign reversal of the longitudinal offset effectively inverted the US East-West axis, cross-correlating local health data with the circadian burden of hypothetical locations on the opposite side of a time zone. We outline the methodology for a correct modelization of the circadian process in the context of US geography.

---

The logic underlying seasonal clock adjustments is ostensibly straightforward: it is a synchronized mechanism designed to align morning social schedules with the seasonal oscillations of sunrise triggered by Earth’s axial tilt at extratropical latitudes (Hudson, 1895). Yet, this simplicity seems to evaporate when modeling the intricate interplay between immutable solar cycles and plastic human activity.

In this report, we establish a framework for the methodologically robust analysis of circadian models under seasonal time shifts. Our critique is prompted by a recent study attributing disease prevalence across United States counties to seasonal clock exposure (Weed and Zeitzer, 2025). Our re-analysis reveals fundamental methodological oversights in that work —most notably, a geographical inversion in which US counties were inadvertently “flipped.” This error resulted in cross-correlating local disease prevalence with the hypothetical circadian burden of locations situated on the opposite side of a time zone. Consequently, the original study’s conclusions currently lack empirical support.

Our analysis focuses on the yearly circadian burden. As previously noted, the effects reported in the original study were negligible —on the order of the model’s own time step— which challenges the validity of associating such minute shifts with bulk societal health outcomes (Martín-Olalla and Mira, 2026). Furthermore, the study appears susceptible to ecological fallacy; it cross-correlates individual-level circadian stressors with aggregate societal outcomes, a known source of epidemiological bias (Erren *et al*., 2026). Our work incorporates a recent correction to the original study.(Annonymous, 2026)

## 1 METHODS

Weed and Zeitzer (2025) utilize a circadian model with one regular schedule: 7am-10pm wake, 9am-5pm work-time in weekdays, and free time weekends. Illumination conditions are fixed to 500 lx at work and to 120 lx in the evening, otherwise determined by ambient light capped at 10 klx.(Weed and Zeitzer, 2025, Figure 1A). Two social clocks are analyzed: the current seasonal clock with two biannual transitions, and a locked clock.

**Figure 1.**
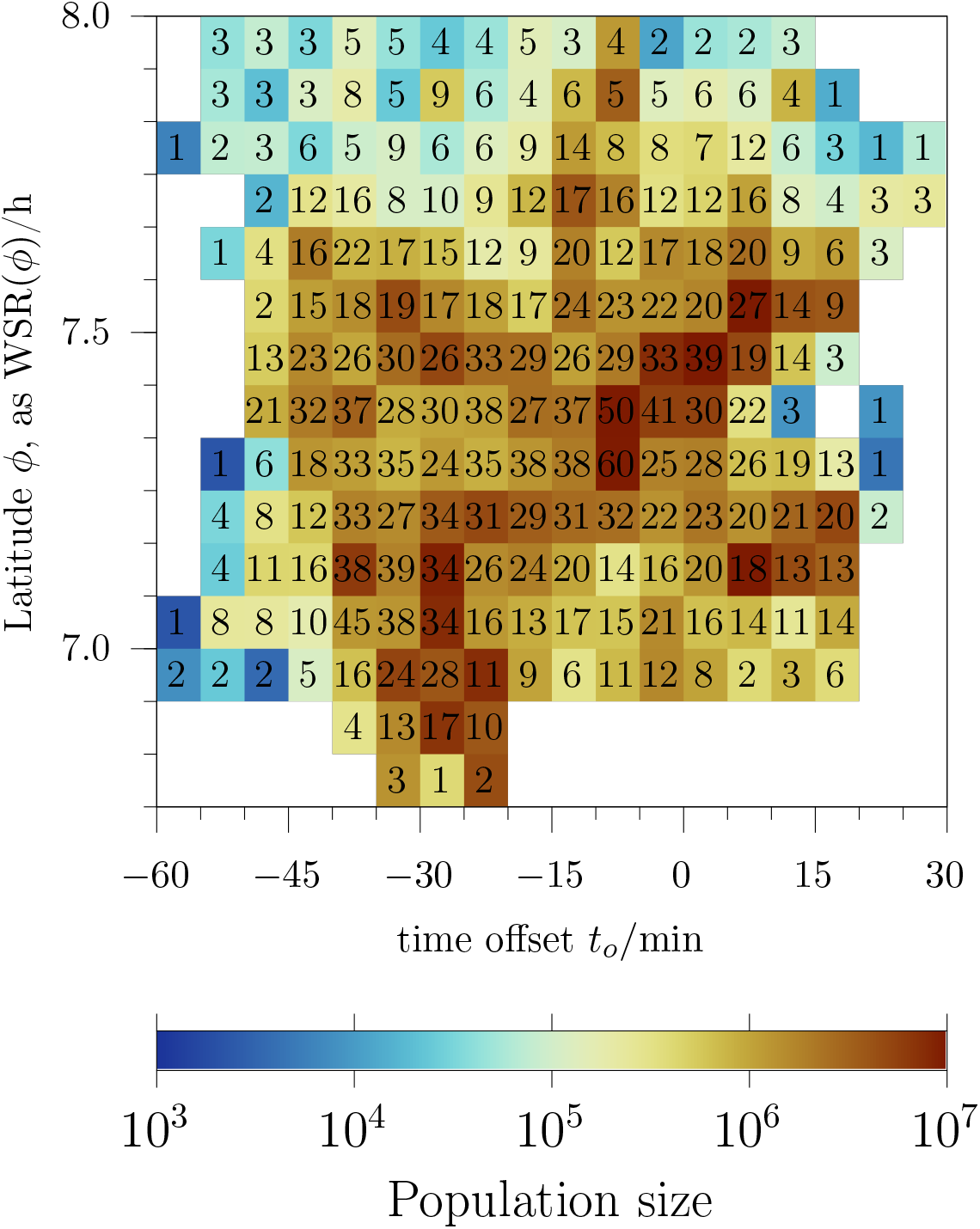
Geographic distribution of US counties. Bin distribution of US counties (*N* = 3108) by centroid time zone offset and latitude, parametrized by winter sunrise time WSR(*ϕ*). Counties are allocated in 207 distinct bins, with 29 bins containing at least 30 counties. See figure 2 for the distribution of centroids and the values of *ϕ*. Palette color is “roma”, see Crameri (2023).

Temporal dynamics are modeled using a 365-day time series with a resolution of *δ* = 5 min. In a scientific context, this series represents mean solar time *t*_*s*_, allow-ing the solar brightness and predefined light conditions to drive the equations for the circadian L-process and P-process uniformly, from which the phase of the circadian clock (the time at core body minimum temperature CBT_min_) is obtained.(Weed and Zeitzer, 2025, Figure 2) Latitude *ϕ* alters solar brightness, sun’s height at noon, and sunrise/sunset times with marked seasonality.

**Figure 2.**
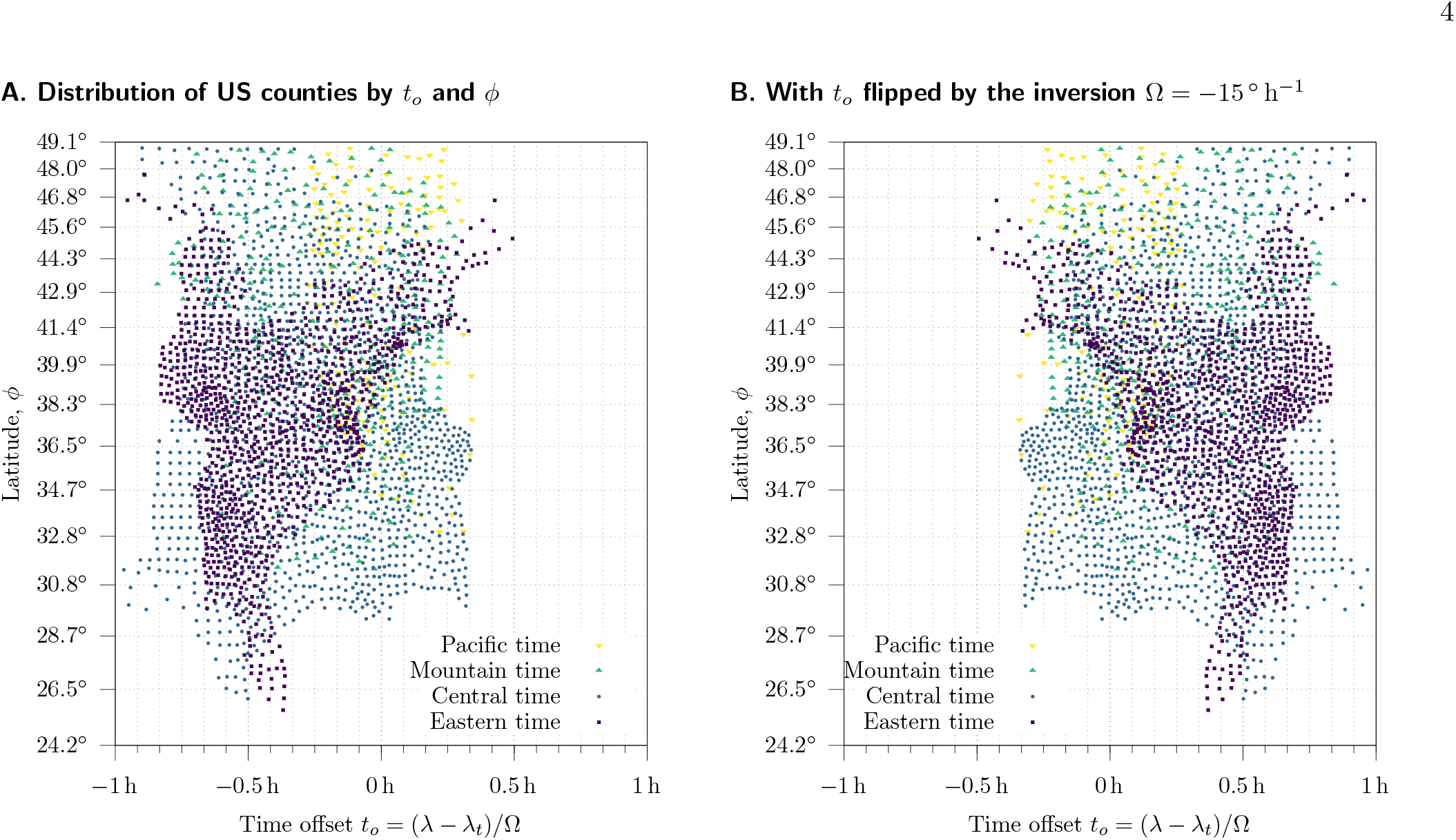
Distribution of US counties by centroid time offset *t*_*o*_ and centroid latitude *ϕ*. The grid matches with the bins in figure 1, which allows to identify bin counts. Distinct US time zones are shown by different shapes and colors. Left: the real distribution with Ω = 15 °h^−1^. Right: the flipped distribution with Ω = −15 °h^−1^ utilized by Weed and Zeitzer (2025).

The boolean conditions that define light diets —for instance, between 9:00 am and 5:00 pm Monday to Friday, the illumination is fixed at *worktime* levels— are set by clock time (social time) *t*_*c*_ and must be mapped to solar time to account for the longitudinal gradient created by time zone standardization. The shift is given by the time offset *t*_*o*_, which represents the longitude *λ* offset relative to the corresponding time meridian *λ*_*t*_, scaled by Earth’s rotation period Ω = 15 ° h^−1^:

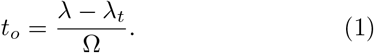

Equation (1) establishes the discrepancy between 12:00 social time and solar noon. West of the time meridian (*λ* − *λ*_*t*_ < 0 and *t*_*o*_ < 0), solar noon occurs after 12:00 social time. Consequently, *t*_*s*_ lags behind *t*_*c*_: for a fixed *t*_*c*_ (e.g. 9:00 am), the corresponding solar time *t*_*s*_ = *t*_*c*_ + *t*_*o*_ decreases westward as *t*_*o*_ becomes more negative, meaning that *t*_*s*_ is progressively earlier. This behavior is inverse to the conventional observation that sunrises occur later in the West.

Within this framework, Standard Time (ST) and Daylight Saving Time (DST) differ simply by a one-hour shift in *t*_*o*_, as the *λ*_*t*_ moves 15° East in the spring and 15° West in the autumn. Consequently, the transition to DST (or to ST) causes *t*_*o*_ to decrease (increase) becoming more (less) negative. The direction of this shift is inverse to the conventional “spring forward, fall back” nomenclature.

Given the model’s temporal resolution, *t*_*o*_ can be partitioned into discrete 5 min bins, ensuring that the underlying boolean conditions remain satisfied within the same time step. Likewise, latitude can be parametrized by the winter sunrise solar time WSR, which acts as a synchronizer of human activity:(Martín-Olalla, 2018, 2019)

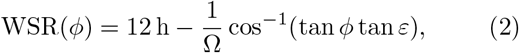

where *ε* = 23.45° is Earth’s axial tilt. Equation (2) approximates sunrise at the solar center and can be reversed. Consequently, partitioning the spatial domain into 5 min intervals of WSR(*ϕ*) is appropriate in this context. Figure 1 illustrates the joint distribution of US counties analyzed in Weed and Zeitzer (2025) grouped by their respective *t*_*o*_ and WSR(*ϕ*) centroid values. Notably, 29 distinct bins contain at least 30 counties, serving as a robust data density metric, or a geographic “h-index” of sorts, for the analysis of societal outcomes.

The original study deviate from this approach in two critical aspects. First, the time series was treated as fixed clock time. In line 44 of the script `computeCircadianShifting.m`,(Weed, 2025) illumination conditions (`computeLux`) were corrected by *t*_*o*_ while the boolean conditions that define light diets remained uniform (lines 50 ff.). This reverses physical reality, where solar conditions are uniform at a given latitude while schedules vary, and introduces a false sense of data density that obscures the discrete nature of the time-series and prevents consistency checks.

Second, a fundamental sign error occurred when in-corporating *t*_*o*_ into lux values. In line 41 of the script, the longitude offset —named `LonOffCenter`, computed in line 103 of `compileCensusData.m`— is divided by Ω = −15 ° h^−1^ —instead of by Ω = 15 ° h^−1^— to compute *t*_*o*_, a variable named `eastWest_hours` in the script. While the script comments correctly state that it is “brighter earlier in the east”, the instruction to “so advance the clock” produced a contradiction: in their model the *t*_*s*_ West of the reference meridian failed to lag behind *t*_*c*_, which is nothing but *t*_*s*_ at the reference meridian. Specifically, at 15° West of the meridian, the brightness belonging to the 10:00 am time step was erroneously assigned to 9:00 am (hence 9:00 am was brighter West of the reference meridian), rather than the 8:00 am brightness required by the longitudinal delay (hence 9:00 am is less bright West of the reference meridian). This error effectively inverted the US East-West axis within the model, see figure 2. Because the original study cross-correlated local disease prevalence of real counties to these geographically “flipped” counterpart, the reported associations are based on a mismatch, rendering its conclusions unsupported.

The daily shifting time Δ_*i*_ is the time difference between two consecutive daily phases Δ_*i*_ = CBT_min_[*i* + 1] − CBT_min_[*i*]. The yearly circadian burden *Y* was computed as *Y* =Σ_*i*_ |Δ_*i*_ |.(Weed and Zeitzer, 2025) We observe that by adopting absolute values, this metric is insensitive to the sign of Δ_*i*_ and is consequently mathematically incapable of differentiating between a stable, regulated circadian phase and a non-regulatory phase that monotonically drifts a comparable cumulative value over a year. In contrast, the standard deviation (SD) of the yearly distribution of Δ_*i*_ provides a more physiologically and statistically grounded alternative, specially for the purpose of correlating circadian stressors with health outcomes, the SD represents a more robust metric.

In our framework, we selected WSR(*ϕ*) values ranging from 6.75 h (*ϕ* = 24.2°) to 8 h (*ϕ* = 49°) while intentionally expanding the analytical scope of *t*_*o*_ from −3 h to 0.5 h. This extended lower bound for *t*_*o*_ was not intended to simulate an extreme “double Daylight Saving Time” scenario; rather, it is parametrized to evaluate conditions corresponding to earlier wake and occupational schedules. Within this methodological approach, *t*_*o*_ effectively operationalizes a macro-social time shift relative to the conventional 9:00 am–5:00 pm baseline schedule.

## 2. RESULTS AND DISCUSSION

Figure 3A displays a heat map of *Y* . On the horizontal axis the reference 0 shows results for the baseline 9:00 am to 5:00 pm at a time meridian. Negative (positive) values represent earlier (later) schedules which can be dynamically operationalized by: (i) moving West (East) within a time zone; (ii) shifting the time meridian East (West) as in transitioning from ST to DST (DST to ST); or (iii) shifting social schedules earlier (later) in absolute time; the horizontal bars between the panels punctuate these shifts, their extension match to figure 1. The burden generally scales with chronotype, increasing from top to bottom panels. Within a chronotype, a horizontal shift toward the left (see the black arrows) generally decreases the burden within the first third of the plot.

**Figure 3.**
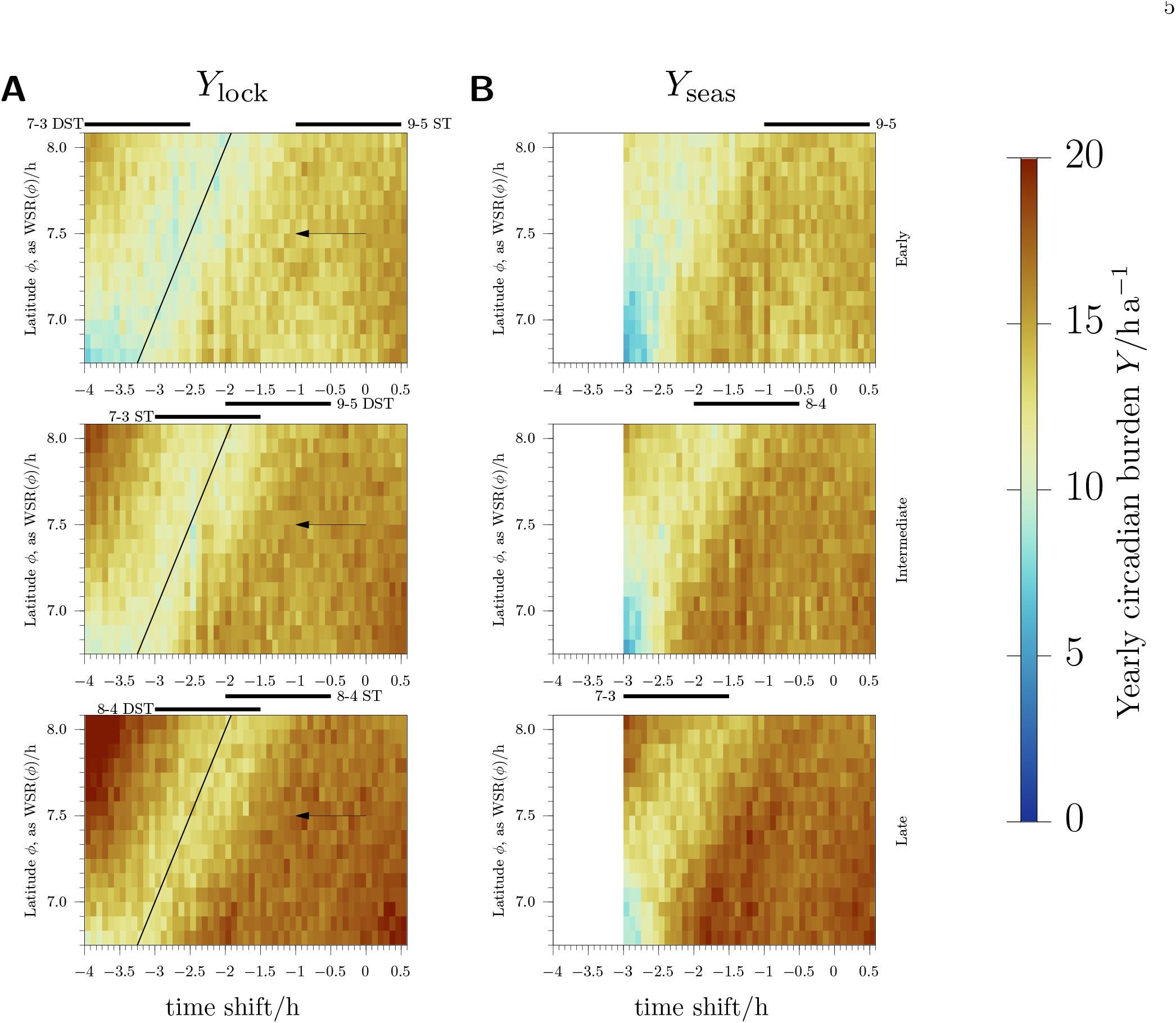
(A) Modeled yearly circadian burden *Y* under locked, non-seasonal clock policies. (B) *Y* under the current seasonal alternating clock policy. Color bar on the right indicate the scale range with higher burdens rendered in red, lower burdens in blue. Vertical panels show results for three chronotypes: early (*τ* = 24 h, top), intermediate (*τ* = 24.2 h, middle), and late (*τ* = 24.4 h, bottom). The time shift is relative to the conventional 9:00 am-5:00 pm baseline schedule used to reproduce the model at a given time meridian; moving horizontally twelve bins (1 h) to the left is equivalent to moving West by 15°in a time zone, moving the time meridian 15°East (i.e., transitioning to DST); or adopting an early 8:00 am-4:00 pm social schedule. Horizontal bars visualize working hours across the extension of the US (see figure 1). In panel A the arrows visualize the shift from ST to DST, and the diagonal line (slope equal to 1) shows the coupling with WSR(*ϕ*).(Martín-Olalla, 2019) Palette color is “roma”, see Crameri (2023).

The burden is low: a total of 16 h per year averages to only 2.63 min per day, or 0.2 % of a day. It is improbable that such marginal variations could yield statistically significant societal health outcomes, given the inherent limitations of the model. Furthermore, a theoretical rationale linking the SD or Σ |Δ_*i*_| —both metrics of the signal variability— to health outcomes remains absent.(Martín-Olalla and Mira, 2026; Weed and Zeitzer, 2025, 2026) Figure 3A reveals a dispersed distribution of *Y* showing perceptible fluctuations for minor changes in *t*_*o*_ and WSR. This dispersion further challenges the validity of *Y* as a reliable predictor of health risks within the current model.

Generally, transitioning from Standard Time (ST) to Daylight Saving Time (DST) schedules decreases the burden, as illustrated by the horizontal thick bars. A region characterized by a comparatively low *Y* (rendered in blue) runs diagonally from SW to NE. The overlaid black line features a unit slope, demonstrating a clear coupling between *Y* (a yearly magnitude) and WSR (a seasonal quantity) within this region: southerners can sustain earlier schedules without a deterioration in *Y* as they seldom find dark morning hours.(Martín-Olalla, 2018, 2019) For the specific geography of the US, a conventional social schedule of *t*_*c*_ = 9:00 am corresponds to approximately 1.5 h after WSR. Consequently, the model suggests that an advance of the social schedule could ameliorate *Y* .

Figure 3B shows *Y* for the current US seasonal clock. It largely reproduces the layout of the locked-clock baseline in figure 3A. Finally While earlier times yield lower *Y*, see the arrows in figure 3A, the biannual clock transitions inflate *Y* immediately following transition dates by design. These two competing effects essentially cancel out, leaving small net differences, see Figure 4A.

**Figure 4.**
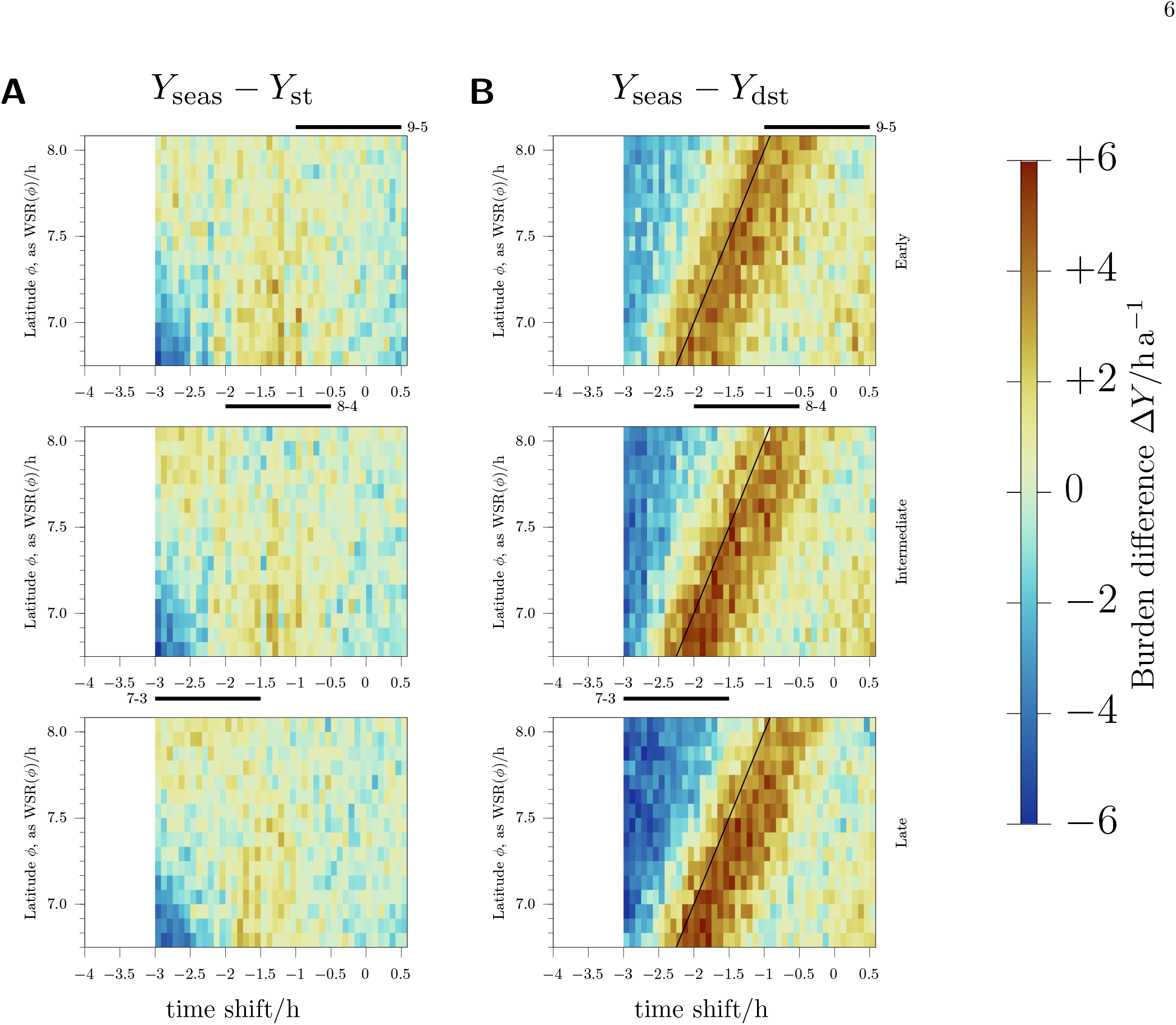
(A) The direct difference *Y*_seas_ *−Y*_lock_. (B) The direct difference *Y*_seas_*− Y*_lock_ with *Y*_lock_ shifted 1 h to the left. Color bar on the right indicate the scale range with positive differences in red, negative differences in blue. Vertical panels show results for three chronotypes: early (*τ* = 24 h, top), intermediate (*τ* = 24.2 h, middle), and late (*τ* = 24.4 h, bottom). The time shift is relative to the conventional 9:00 am-5:00 pm baseline schedule used to reproduce the model at a given time meridian; moving horizontally twelve bins (1 h) to the left is equivalent to moving West by 15°in a time zone, or adopting an early 8:00 am-4:00 pm social schedule. Horizontal bars visualize working hours across the extension of the US (see figure 1).In panel B the diagonal line (slope equal to 1) shows the coupling with WSR(*ϕ*).(Martín-Olalla, 2019) Palette color is “roma”, see Crameri (2023).

Figure 4B compares the current seasonal clock with the DST clock, obtained by shifting *Y*_lock_ 1 h to the left. Alternatively, a 1 h displacement left to right in Figure 3A is used. In this scenario the two underlying mechanics reinforce one another: shifting to the right in figure 3A increases *Y*, and the biannual transitions further inflates *Y*, which results in positive Δ*Y* in figure 4B. This panel is dominated by a South West to North East band in red (meaning conditions worsened by seasonal clocks) which recalls the diagonal band in figure 3A.

We do not conclude from this specific interaction that permanent DST is superior to permanent ST or the current seasonal clocks. First, individuals aligned with earlier schedules would experience a distinct deterioration of *Y* . Second, because *Y* is fundamentally low and behaves unsmoothly across narrow parameter windows, it cannot serve as a robust deciding metric for public policy. Notwithstading these limitations, it may provide a subtle quantitative clue regarding the current preferences for DST expressed by polls.(Rubin, 2023)

The ultimate takeaway of this methodological critique is straightforward: in a scientific context, one must always anchor the analysis to physical reality. Solar conditions must be taken as given; it is the flexible adaptation of social schedules that dictates the alignment. Socially speaking, the current seasonal clock must be understood as an adaptive compromise between two extreme choices.(Martín-Olalla and Mira, 2025)

## Data Availability

All data produced in the present study are available upon reasonable request to the authors

## ACKNOWLEDGMENTS

The authors acknowledge the use of an Artificial Intelligence large language model, Gemini, to refine the draft manuscript for grammar and clarity. Gemini was made available to the authors through a collaborative initiative between Google and Universidad de Sevilla. Mat-Lab©MathWorks 2025b was used to compute the circadian model. MatLab was made available to the authors through a collaborative initiative between MathWorks and Universidad de Sevilla.

## AUTHOR CONTRIBUTIONS

All authors contributed equally to this work.

